# Detection of Hypervirulent *Klebsiella pneumoniae* from Clinical Samples in Tamil Nadu

**DOI:** 10.1101/2023.02.19.23286158

**Authors:** Thamaraiselvan Shanthini, Prasanth Manohar, Xiaoting Hua, Sebastian Leptihn, Ramesh Nachimuthu

## Abstract

*Klebsiella* pneumoniae is one of the significant opportunistic pathogens which cause both nosocomial and community-acquired infections. This study focuses on the molecular characterization of hypervirulent *K. pneumoniae* isolated from clinical samples in Tamil Nadu. A total of 30 clinical *K. pneumoniae* isolates were collected from the diagnostic centres located in Trichy, Madurai, and Chennai. On subjected to the antibiotic susceptibility testing, it was found that 73% were multi-drug resistant. The string test showed that 46.6% of the isolates were hypermucoviscous (HMV). Molecular studies revealed the absence of *mcr (1-5)* genes. The isolates belonging to capsular types K1 (n=5), K2 (n=6) and K5 (n=2) were detected. Virulence genes namely *rmpA* (n=5), aerobactin (n=8), and *KfuBC* (n=8) were identified using multiplex polymerase chain reaction. Through molecular studies, 22 isolates were found to be classical *K. pneumoniae* (cKP) and 8 were hypervirulent *K. pneumoniae* (hvKP). The genome sequencing of five isolates revealed that the strains belong to ST86 and ST23. *In vivo* studies using *Galleria mellonella* showed that HMV-hvKP strains were highly pathogenic among the hvKP strains and the non-K1/K2 and K2 strains were pathogenic among the cKP strains. Our data revealed the emergence of colistin-resistant hvKP strains in Tamil Nadu, India.

## Introduction

*K. pneumoniae* is one of the distinguished members of the *Enterobacteriaceae* family known to cause pneumoniae, urinary tract infection (UTI), sepsis, meningitis, soft tissue infection and pyogenic liver abscesses (PLA).^1^ The accretion in antibiotic resistance among Gram-negative bacteria makes most of the definite treatment defenceless. The progression of resistance against the last-resort antibiotics is increasingly reported in *K. pneumoniae;* which eventually becomes highly challenging for clinicians to combat as it is sprouting with hypervirulence.^2,3^ At first, it was observed that the hypervirulent strains were mostly susceptible to available antibiotics. In recent years, there have been perpetual reports worldwide on the emergence of hypervirulence and hypermucoviscous phenotypes convergence with a high level of resistance.^4^ The emerging combo of hypervirulence and resistance such as carbapenem-resistant *K. pneumoniae* (CRhvKP), hypervirulent colistin-resistant *K. pneumoniae* (ColRhvKP) and hypervirulent tigecycline-resistant *K. pneumoniae* (TighvKP) needs immediate attention. Since they are likely to cause community-acquired infections and can infect immunosuppressed patients including young and healthy individuals.^5,6^ Recent reports on its association with the plasmid-mediated resistance gene, *mcr-1* create panic among clinicians. It could quickly transfer its resistance to different species (horizontal gene transfer), and the possibility of virulence transfer is high.^7^ The hypervirulent (hvKP) strains are mainly associated with hypermucoviscous phenotype, which signs positive for string test (hmKP), explicitly K1 and K2 capsular serotypes are known to elevate the hyper capsule production and aerobactin, the important siderophore. This dominant siderophore is considered one of the essential factors for virulence in bacteria.^8^ Recent reports found that the hypervirulent strain was also found independently from the hypermucoviscous phenotype. In some cases, the hypermucoviscous phenotype is also found to be independent of capsule production.^9^ There is no standard definition and marker to characterize hypervirulent *K. pneumoniae* and its link with hypermucoviscous *K. pneumoniae* strains.^10^ The prevalence of hypervirulent *K. pneumoniae* (hvKP) strains is in Asia; however, there is a comprehensive indication of its dissemination, which needs to be curbed.^11^ The use of *Galleria mellonella* (wax moth larvae) is found to be an effective alternative model to study bacterial pathogenesis with significant advantages including the innate immune response of wax moth larvae (non-vertebrate) is similar to vertebrates.^12^

This study aimed to screen for colistin- and tigecycline-resistant *K. pneumoniae* among the clinical isolates and to identify the prevalence of hypervirulent *K. pneumoniae* in the study region. Further, the hvKP isolates were characterized at the genomic level and clonal similarities were determined. The severity of the hvKP strains was studied using *G. mellonella* as a model organism.

## Materials and Methods

### Collection and identification of clinical strains of K. pneumoniae

A total of 30 clinical isolates of *K. pneumoniae* were collected from three different diagnostic centres located in Chennai, Trichy and Madurai in Tamil Nadu, India. The isolates were cultured from various clinical sources including urine, blood, sputum, pus, and cerebral spinal fluid (CSF). The isolates were transferred under cold conditions to the Antibiotic resistance and Phage therapy laboratory at VIT, Vellore. All 30 *K. pneumoniae* isolates were sub-cultured on MacConkey and HiCrome™ *Klebsiella* selective medium (Himedia, India). Further, the *Klebsiella* species was confirmed using VITEK 2 identification system (bioMerieux). This study was conducted as per the ethical guidelines issued by the Institutional Ethical Committee for the studies on Human subjects (IECH) VIT/IECH/004/Jan28.2015.

### Antibiotic susceptibility testing

All 30 *K. pneumoniae* were subjected to disc diffusion by the Kirby-Bauer method. The following antibiotic discs (Himedia, India) were used for testing the sensitive pattern: Amikacin (AK, 30 mcg), Gentamicin (GT, 10 mcg) from aminoglycosides; Amoxyclav (Amoxicillin/clavulanic acid AMC, 10 mcg) from beta-lactamase inhibitor; Cefotaxime (CX, 30 mcg), Cefepime (CF, 30 mcg) third and fourth generation of cephalosporin; Meropenem (MR, 10mcg), Ertapenem (ER, 10 mcg) from carbapenem was performed according to the Clinical Laboratory and Standards Institute (CLSI) guidelines (CLSI, 2018). Colistin discs were not used because their poor diffusion on agar may produce a defective result. Besides, there is no legitimate interpretation of the breakpoint for colistin disc diffusion in EUCAST and CLSI guidelines.^13,14^ Hence, minimum inhibitory concentration (MIC) was performed using broth dilution for colistin and tigecycline according to CLSI guidelines. The results were interpreted using EUCAST guidelines for colistin (EUCAST, 2018) and FDA guidelines for tigecycline (FDA, 2010).^13,15^ *E. coli* ATCC 25822 and *P. aeruginosa* ATCC 27853 were used as control strains.

### String test

To identify, hypermucoviscous *K. pneumoniae*, the strains were sub-cultured on 5% sheep blood agar. The mucoviscosity was assessed by stretching a single bacterial colony using a loop. It is considered a positive string test when the bacterial colony was touched with a sterile loop and stretched. The formation of viscous string >5 mm was considered hypermucoviscous, whereas the strains that showed a viscous string of <5 mm were considered non-hypermucoviscous.^16^

### Molecular characterization of virulence and resistance

Genomic DNA was isolated using the boiling method. In brief, the overnight culture was spun at 7000 × g for 15 min. The aqueous layer was discarded, and the pellet was resuspended with 100 μL of sterile distilled water. The bacterial suspension was centrifuged at 7000 × g for 10 min, and to the pellet, 50 μL of sterile distilled water was added. The pellet was incubated in the water bath at 100°C for 15 min. Further, it was allowed to cool for 5 min and centrifuged at 7000 × g for 5 min. The aqueous layer containing DNA was aliquoted into a fresh tube and stored at -20°C. For PCR analysis, 2 μL of template DNA was used.^17^ All the phenotypically identified colistin-resistant *K. pneumoniae* isolates were screened for the presence of mobile colistin-resistance genes (*mcr-*1 to *mcr-*5) using the multiplex PCR method as described earlier.^18^ The primers sequence was provided in table S1.

In the case of the integron study, class-1, class-2, and class-3 integrons were screened in all the colistin-resistant *K. pneumoniae* isolates using the integrase genes, *IntI1, IntI2 and IntI3*. The class 1 integron gene cassette regions were screened using *5’CS* and *3’CS*, and the size of the variable region was detected.^19^ The primers sequence was provided in table S2.

For virulence screening, we subjected all the *K. pneumoniae* isolates to screen for the capsular serotypes, namely K1, K2, and K5, which are highly associated with invasive infection. In addition, virulence genes, *rmpA, aerobactin*, and *kfuBC* were screened. The primers and PCR conditions were described earlier.^20,21,22^ The primers sequence was provided in tables S3 and S4.

### Genome analysis of hypervirulent K. pneumoniae

The bacterial strains were cultured overnight in LB broth at 37°C. The bacterial genomic DNA was extracted using the HiPurA Bacterial Genomic DNA Purification kit (Himedia, India). The genome was sequenced on an Illumina HiSeq4000 platform using a paired-end 2 × 150 base pair protocol by Macrogen Inc. (South Korea). Derived raw reads were *de novo* assembled using Shovill 1.0.9 software. The generated contig sequences were annotated by Prokka v1.13. The core alignment was generated via panaroo v1.1.2. And the phylogenetic tree was constructed by iqtree v2.0.3. And the tree was plotted using the R package ggtree.

### In vivo studies using G. mellonella

For *in vivo* studies, *G. mellonella* was obtained from the Department of Entomology, University of Agricultural Sciences, Gandhi Krishi Vignana Kendra, Bengaluru, India. The moths were grown at 35°C and once the worms reached their late larval stage, they were used for the study. A total of four hvKP (KP1, KP2, KP3 and KP4) and three cKP (KP5, KP6 and KP7) isolates were selected. The bacterial culture was adjusted to 10^4^, 10^5^, and 10^6^ CFU/mL using phosphate buffer saline (PBS) for intrahemocoelic injection. The selected larvae were surface disinfected with 70% (v/v) ethanol and injected with 20 μL of bacterial suspension using the syringe into the last left pro-leg. At least 10 larvae were used in each group and larvae injected with PBS served as a control. The study groups consist of larvae infected with hvKP and cKP strains. The infected larvae were considered infected/dead when they were turned black or without movement. The larvae were counted for their survival every 12 hours for five days and the time of death was recorded.^6^

## Results

### Identification and distribution of colistin resistance in K. pneumoniae isolates

A total of 30 clinical isolates were identified to be *K. pneumoniae*, of which 22/30 (73%) isolates were found to be multidrug-resistant by disc diffusion. The complete list of isolates and their clinical source is provided in table S5. The highest resistance rate was observed in amikacin (100%) followed by gentamicin (83%), amoxiclav (76%), cefotaxime (63%) and cefepime (56%) and the lowest resistance was observed in meropenem (16%) and ertapenem (6%). In this study, eight hvKP and 22 cKP isolates were identified. Among the hvKP isolates, 4/8 (50%) were MDR, and 18/22 (81.8%) were MDR in cKP isolates. It was found that 27/30 (90%) isolates were resistant to colistin, and all the isolates were sensitive to tigecycline by MIC. Among the hvKP isolates, 7/8 (87.5%) were resistant to colistin, and in cKP isolates, 20/22 (90.9%) were resistant to colistin.

### Characterization of hypermucoviscosity in K. pneumoniae isolates

Among the 30 *K. pneumoniae*, 14 (46.6%) isolates were positive for the string test. The isolates which form the string of >5 mm were considered HMV *K. pneumoniae*. Among the hvKP isolates, 5/8 (62.5%) had an HMV phenotype, whereas 8/22 (36.36%) were HMV in cKP isolates. On genotypic characterization, it was found that 5/14 string positive isolates carried the *rmpA* gene and all the five isolates were hvKP. The 5 hvKP isolates carried the *rmp*A gene, which is responsible for the mucoviscosity in *K. pneumoniae*, and the string-positive cKP isolates did not carry the *rmp*A gene.

### Genotypic characterization of virulence genes in K. pneumoniae isolates

On screening for virulence genes, it was found that five isolates carried *rmpA*, eight had *aerobactin* and eight had *kfuBC*. The capsular typing results showed that five isolates were K1, six isolates were K2 and two isolates were K5. The presence of capsular typing and virulence genes in combinations were found as String-K1-Aerobactin-*rmpA-KfuBC* (n=2), String-K2-Aerobactin-*rmpA-KfuBC* (n=1), String-K2-Aerobactin-*rmpA* (n=1), String-K2 (n=1), String-K1-*rmpA* (n=1), String-*rmpA* (n=1), K1-Aerobactin (n=1), K1 alone (n=1), K5-*KfuBC* (n=2), K2-Aerobactin-*KfuBC* (n=2), K2-*KfuBC* (n=1). The capsular typing and the virulence genes were listed in table 1.

**Table 1:** The distribution of virulence genes within different capsular serotypes among the colistin-resistant *K. pneumoniae*

| S.No. | Organism ID | Capsular serotypes | Aerobactin | *rmpA* | String test | KfuBC | Biofilm | HMV KP/non-HMV KP | hvKP/cKP |
| --- | --- | --- | --- | --- | --- | --- | --- | --- | --- |
| 1 | KP 1 | K1 | + | + | + | + | Weak | HMVKP | hvKP |
| 2 | KP 3 | K1 | + | - | - | - | Strong | Non-HMV KP | hvKP |
| 3 | KP 5 | K1 | - | - | - | - | Moderate | Non-HMV KP | cKP |
| 4 | KP 7 | K5 | - | - | - | + | Strong | Non-HMV KP | cKP |
| 5 | KP 2 | K2 | + | + | + | + | No | HMV KP | hvKP |
| 6 | KP 4 | K2 | + | - | - | + | Strong | Non-HMV KP | hvKP |
| 7 | KP 6 | K2 | - | - | - | + | Strong | Non-HMV KP | cKP |
| 8 | KP 8 | K1 | + | + | + | - | Moderate | HMV KP | hvKP |
| 9 | KP 9 | K2 | + | - | - | + | Moderate | Non-HMV KP | hvKP |
| 10 | KP 10 | K5 | - | - | - | + | Strong | Non-HMV KP | cKP |
| 11 | KP 11 | K2 | - | - | + | - | Moderate | HMV KP | cKP |
| 12 | KP 12 | K2 | + | + | + | - | Weak | HMV KP | hvKP |
| 13 | KP 13 | K1 | + | + | + | + | Moderate | HMV KP | hvKP |

**Table 2:**
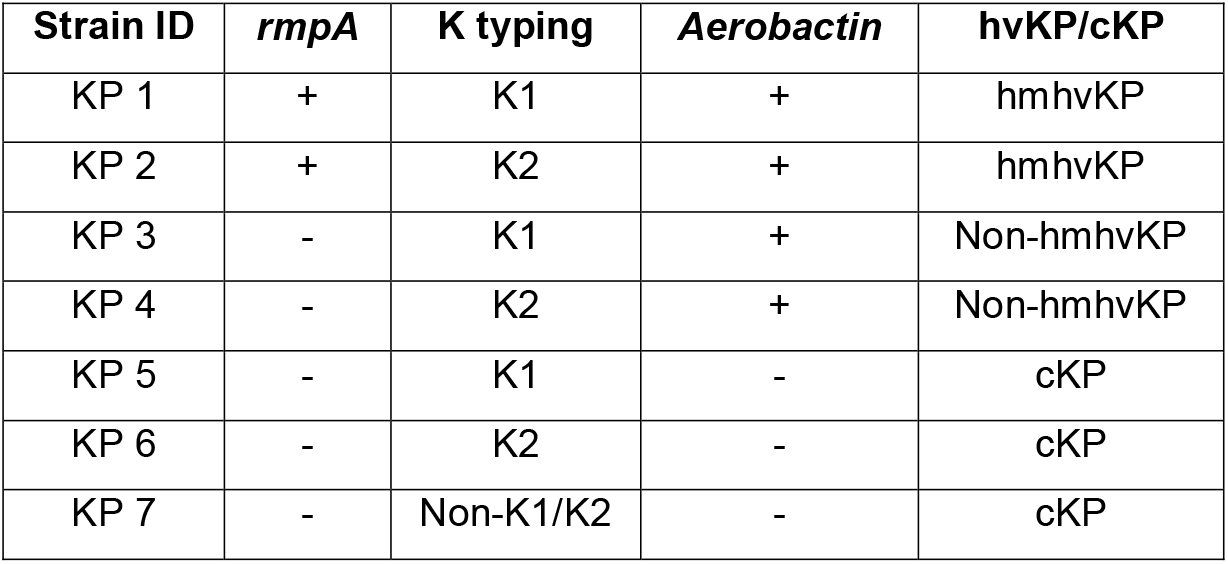
The list of hypervirulent (hvKP) and classic (cKP) *K. pneumoniae* isolates used to study the pathogenesis in the *Galleria mellonella* model.

### Screening of integron and mcr genes in K. pneumoniae isolates

All 27 colistin-resistant *K. pneumoniae* isolates were subjected to class 1, class 2, and class 3 integron genes along with their variable regions. We found that 9/27 (33.3%) of the isolates had class 1 integron but none of the isolates carried gene cassette. Interestingly, none of the isolates detected *mcr* (1-5) genes.

### Genome analysis

A total of five colistin-resistant hvKP strains (KP1, KP2, KP8, KP9 and KP12) were subjected to genome sequencing and analysis (Fig. S1). The five strains were divided into two clusters (Fig.1). Among them, KP9 and KP2 belonged to ST86 and harboured *bla*_SHV-208_, *fosA_gen, oqxA*11 and *oqxB*19. Meanwhile, KP1, KP8 and KP12 formed another cluster (ST23) and harboured *bla*_SHV-190_, *fosA*6, *oqxA*5 and *oqxB*12. The sequence assemblies were deposited under the BioProject: PRJNA838492.

**Figure 1:**
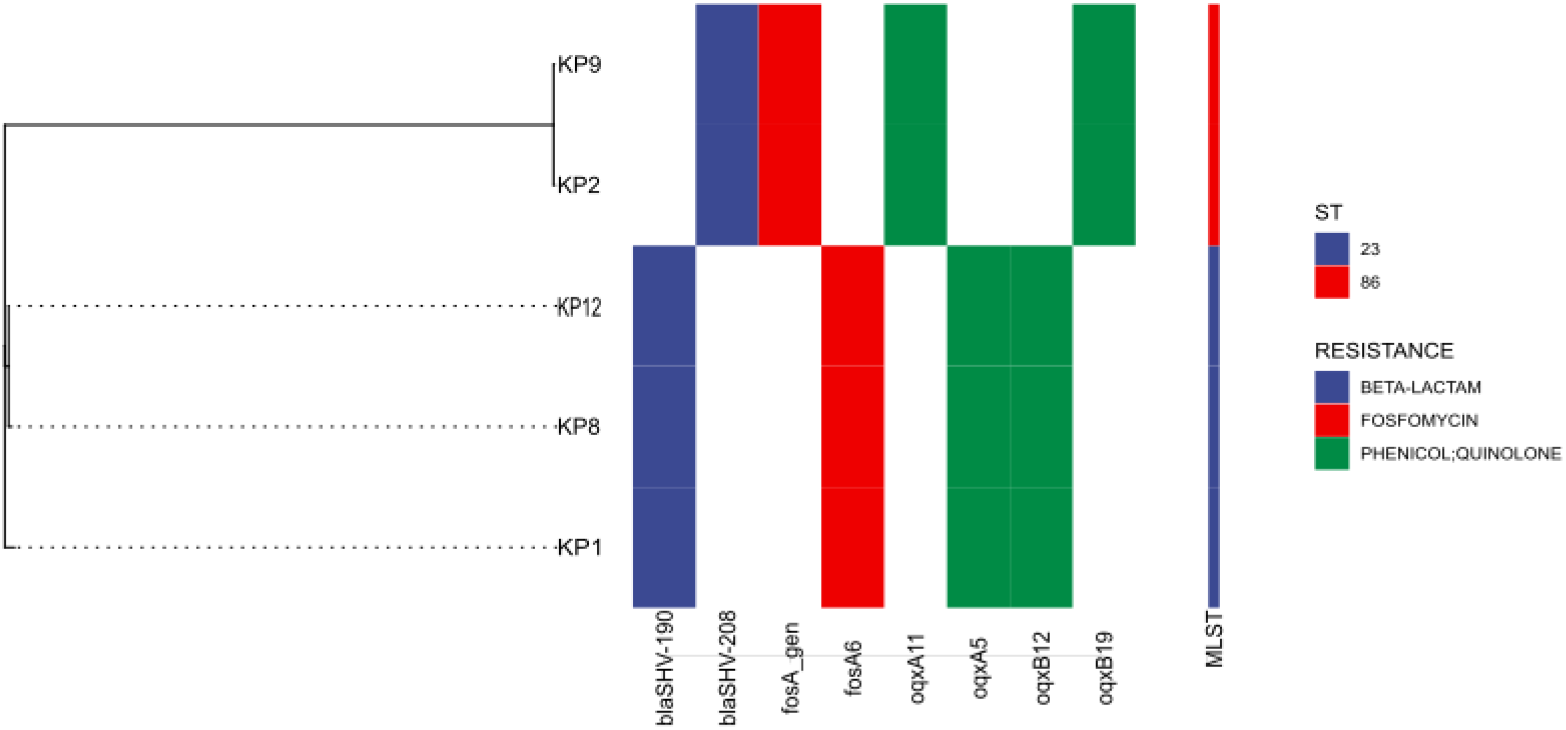
The phylogenetic analysis of the five *K. pneumoniae* isolates. The five strains of colistin-resistant hvKP isolates were sequenced (WGS) and based on MLST it is grouped into two clusters, KP9 and KP2 belonged to ST86, and KP1, KP8 and KP12 formed the ST23 cluster. All the isolates were found to harbour resistance-related genes.

**Figure 2:**
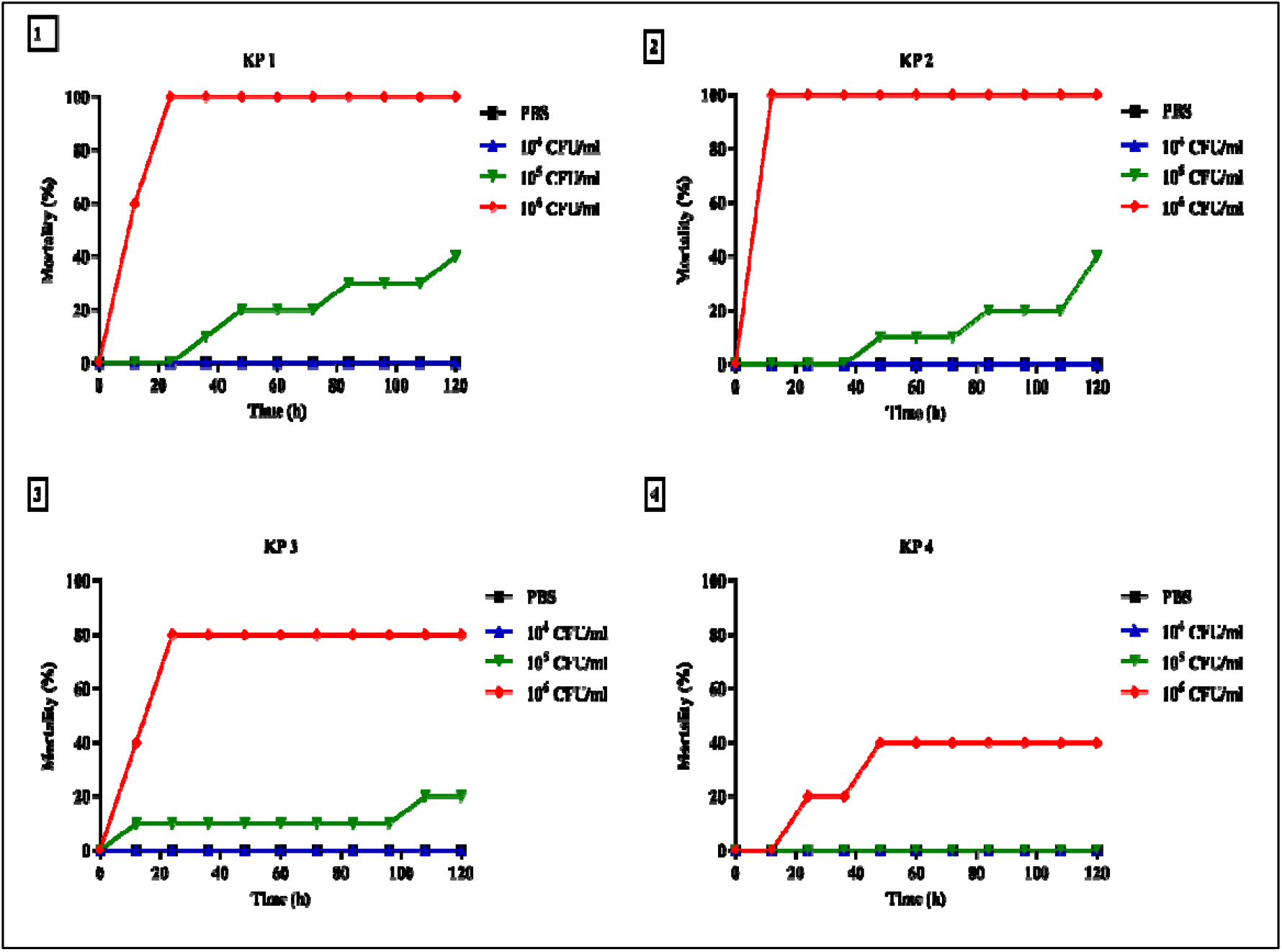
The mortality of *G. mellonella* infected with varying concentrations (1×10^4^, 1×10^5^ and 1×10^6^ CFU/mL) of hypervirulent *K. pneumoniae* (hvKP) strains. The graph represents the % of dead larvae infected with hvKP strains.

There was a spot on the infection site at an inoculum of 1×10^4^ CFU/mL, but no larvae were dead in cKP strains. In contrast, when injected with an inoculum of 1×10^5^ CFU/mL, there was melanization in the larvae, and the capsular K typing positive cKP strains KP5 and KP6 attained 30% and 100% mortality at 120 hours. But the non-K1/K2/K5 cKP strainKP7 had 30% mortality at 120 hours. At an inoculum of 1×10^6^ CFU/mL, the capsular K typing cKP strains KP5 and KP6 attained 40% and 100% mortality at 24 and 12 hours, whereas the non-K1/K2/K5 cKP strains, namely KP7 attained 100% mortality in 12 hours (Fig.3).

**Figure 3:**
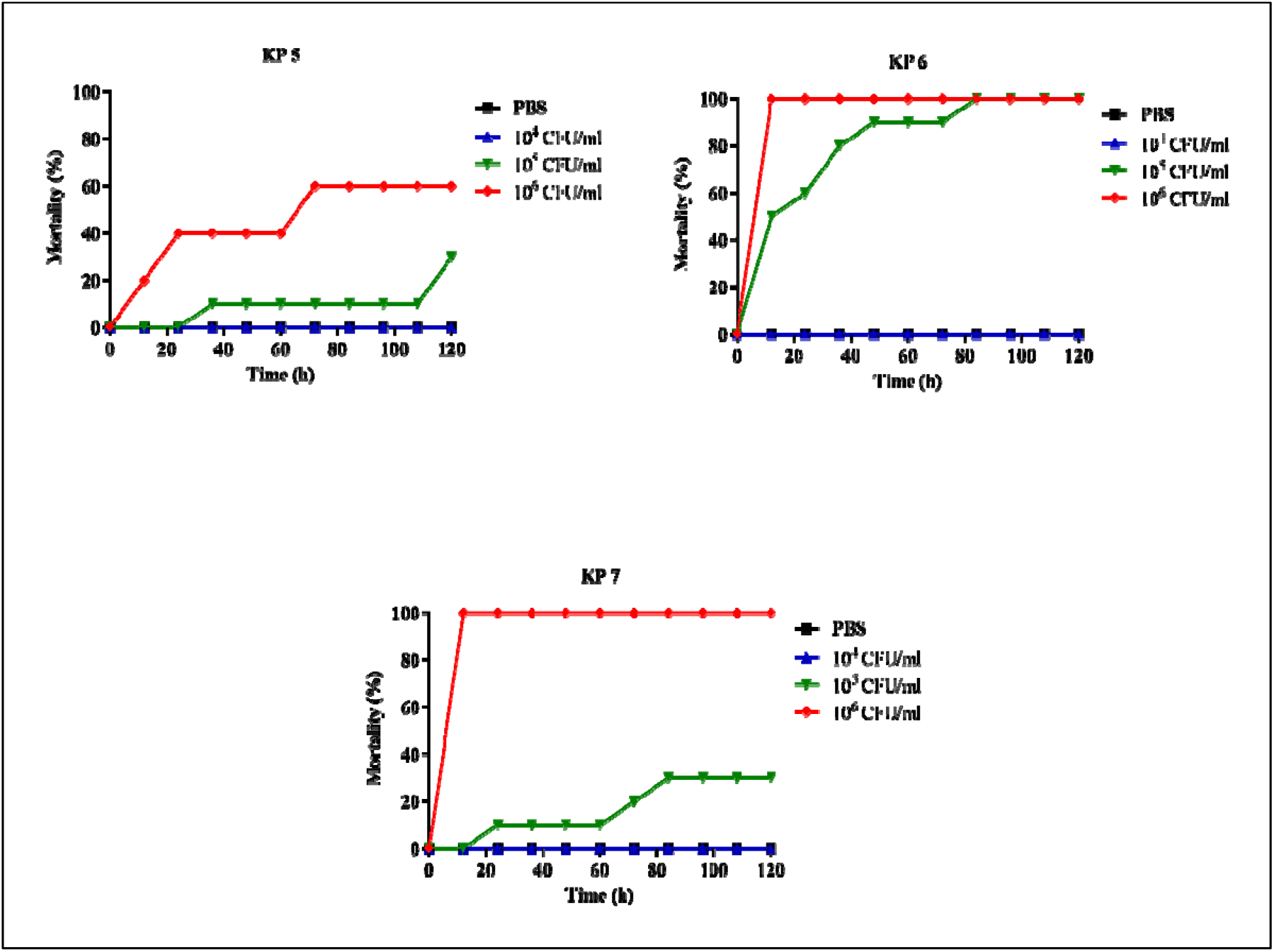
The mortality of *G. mellonella* infected with varying concentrations (1×10^4^, 1×10^5^ and 1×10^6^ CFU/mL) of classical *K. pneumoniae* (cKP) strains. The graph represents the % of dead larvae infected with cKP strains.

It shows that in the case of capsular K typing cKP strains, the KP6 with K2 serotype is highly pathogenic compared to the K1 positive KP5 cKP strain. In contrast, the non-K1/K2 strain, namely, KP7, was also found to be pathogenic at higher concentrations, which indicates that any infection with non-K1/K2/K5 strains can be lethal.

## Discussion

Among the GNB, *E. coli* and *K. pneumoniae* play a pivotal role in disseminating nosocomial infections, and they evolve rapidly by adapting to different environments.^23^ In India, *K. pneumoniae* is found to cause UTI, bloodstream infections, meningitis, pneumonia, and endophthalmitis in neonates. The startling case of *Klebsiella* infection dynamics in Indian hospitals and communities is increasingly reported.^24,25^ *K. pneumoniae* is considered a deadly superbug and it is emerging with hypervirulence properties. It is important to explore the molecular characteristics of the emerging strains of the new hypervirulent *K. pneumoniae*. Hence, this study explores the resistance and virulence characteristics of *K. pneumoniae*.

There is an increase in cases of colistin resistance in Eastern India, where they reported 24 cases of colistin-resistant *K. pneumonia* infection in immunocompromised patients.^26^ Likewise, a report from Christian Medical College, Tamil Nadu, reported eight colistin-resistant *K. pneumonia* strains from patients with bloodstream infections and found there is a steep increase in colistin resistance infections among the GNB, particularly nosocomial *K. pneumoniae*.^27^ In this study, the distribution of colistin resistance in *K. pneumoniae* was very high up to 90%. But *mcr* (1-5) genes were not detected, which shows a lack of plasmid-mediated resistance, which is similar to our previous report.^28^ The colistin resistance could be due to chromosomal-mediated such as alteration in the LPS target, high mucous production, or biofilm formation.^29^ Through genome sequencing, no unique alternations were identified in colistin-resistant isolates. In certain strains, the resistance was due to chromosomal-mediated; namely, *mgrB* inactivation, alteration, or upregulation in the two-component regulatory system, and in some cases, the resistance was due to the active efflux pumps.^28,30^

All the *K. pneumoniae* isolates used in this study were sensitive to tigecycline. It supports the rare occurrence of tigecycline resistance in our regions and the earlier study where the tigecycline resistance in *K. pneumoniae* was found to be lesser than in other antibiotics.^31^ On the contrary, there is an emerging tigecycline resistance in *K. pneumoniae*.^32^ As stated earlier, the emerging strains of MDR, XDR and PDR *K. pneumoniae* strains were challenging to treat with the last resort antibiotics.^33^

Integrons are one of the critical mobile genetic elements that are highly responsible for the speedy transfer of antibiotic resistance.^34^ The higher prevalence of integrons in the colistin-resistant *K. pneumoniae* is alarming as they could transfer the resistance to other bacteria. It is imperative to study integrons in depth to figure out their stint in radiating antibiotic resistance. In this study, the prevalence of class 1 integrons was found to be 33.3% in the colistin-resistant *K. pneumoniae* isolates, which is similar to the previous report, where they reported 41.6% (10/24) of class 1 integrons in colistin-resistant *K. pneumoniae* isolates. There is an alarming prevalence of integrons in the nosocomial setting. It would be highly dangerous as it might be challenging to curb the spread if not taken properly measures and impede antibiotic therapy.^28^ Also, it agrees with another report, which showed that the availability of class 1 integron in the MDR strains could have an impact on the dispersal of clustered resistance genes.^35^

The detection of hypermucoviscous (HMV) phenotype is feasible and can be easily performed in any microbiology laboratory. It could help to diagnose the HMV strains earlier as most of the HMV *K. pneumoniae* strains are known to cause a liver abscess. It is worrisome that specific *K. pneumoniae*, namely, HMV-hvKP, can cause metastatic spread in infected patients.^36^ In this study, a total of 14 HMV *K. pneumoniae* isolates were identified using a string test, of which 54% (12/22) were resistant to colistin. In a similar study, 22.8% (84/369) of the *K. pneumoniae* HMV phenotype was detected using a string test.^37^ It should be noted that there is a high prevalence of HMV *K. pneumoniae* among the clinical isolates. In this study, the association of HMV *K. pneumoniae* that are resistant to colistin was reported for the first time in South India. Interestingly, genome analysis showed the presence of hvKP ST23 and ST86 strains in the study region. Though ST23 is previously reported in the study region, the presence of hvKP ST86 is first reported in this study.^38^

There is a shred of profound evidence that the HMV *K. pneumoniae* was associated with hypervirulence. Still, there is no concluding evidence to define a strain as hvKP on its association with the HMV.^39^ A study by Fang *et al*., reported that hypervirulence is defined by HMV phenotype using a string test.^40^ In contrast, another study suggested that the HMV alone cannot define a strain as hvKP, whereas other virulence factors such as aerobactin or capsular typing K1/K2 are needed.^3^ In this study, the HMV was observed in five hvKP strains and nine HMV wherein cKP strains. The HMV in cKP strains was reported in another study.^41^ Our study also highlights the presence of non-HMV hvKP strains as reported earlier.^42^ Our study insists on the necessity to screen for hypervirulence in the non-HMV isolates of *K. pneumoniae*.

There is more misinterpretation about the identification of hvKP strains. The detailed insight at the molecular level to study the virulence genes such as *rmpA* and aerobactin, which plays a vital role in hvKP strains as virulence inducers, could help in identification. These virulence genes are involved in increasing mucous production and help the pathogens to acquire iron from the host for their survival. The role of capsular serotype K1/K2 is also highly associated with hvKP strains, but recent reports on hvKP strains were also found in non-K1/K2 serotypes.^43^ Accordingly, in this study, the prevalence of virulence genes was found in 14/30 (46.6%) *K. pneumoniae* isolates. The 11 different combinations of virulence and capsular types found among the hvKP and cKP strains were as follows, String-K1-Aerobactin-*rmpA-KfuBC* (n=2), String-K2-Aerobactin-*rmpA-KfuBC* (n=1), String-K2-Aerobactin-*rmpA* (n=1), String-K2 (n=1), String-K1-*rmpA* (n=1), String-*rmpA* (n=1), K1-Aerobactin (n=1), K1 alone (n=1), K5-*KfuBC* (n=2), K2-Aerobactin-*KfuBC* (n=2), K2-*KfuBC* (n=1). Among the eight isolates, seven were resistant to colistin, and none of them was resistant to tigecycline. Another study from Tamil Nadu also reported carbapenem-resistant hvKP strains.^44^ Likewise, hvKP strains among colistin-resistant *K. pneumoniae* and its emanation in Tamil Nadu were first reported in this study. Also, it is the first study to report colistin-resistant hvKP strains in India to the best of our knowledge. It shows the wide distribution of hvKP strains in India, and in-depth surveillance is necessary. Clinically, the combination of colistin resistance and hvKP is highly challenging to treat and may lead to a high mortality rate. Similar reports on hvKP with colistin resistance were reported in China, where they identified five colistin-resistant hvKP strains from the blood samples collected from five male patients.^6^ A recent report from Taiwan showed the presence of tigecycline-resistant hvKP strains and explored its resistance mechanism.^45^

On the other hand, K-typing alone or the presence of the virulence genes in the chromosome alone is sufficient to characterize a strain to be cKP.^46^ In this study, among the 22 cKP strains, about seven strains had either K1/K2/K5 serotypes or any of the virulence genes, namely *rmpA, kfuBC* or aerobactin gene. The cKP strains are highly associated with multi-drug resistance among *K. pneumoniae*, and in this study, among the cKP strains, 66.6% were MDR. The prevalence of cKP strains was mostly in the western countries,^46^ whereas in our study, the prevalence of the cKP strain was high compared to hvKP strains. Attention is necessary against the newly emerging hvKP strains as they are prevalent in Asia. However, there are minimal studies and reports about hvKP strains in India, indicating either the infections due to hvKP are rare or no monitoring due to lesser knowledge. The dissemination of colistin-resistant hvKP strains reported in this study will alert clinicians and researchers to take immediate steps to control its spread.

Further, *G. mellonella* was used as an infection model to study the pathogenicity of hvKP and cKP strains with various combinations of virulence genes and capsular serotypes. It showed that the colistin-resistant HMV-hvKP strains were highly virulent compared with non-HMV hvKP strains in this study. In the case of cKP strains, the strain positive for K2 serotyping and the non-K1/K2 strain were found to be highly pathogenic when compared to the K1 *K. pneumoniae* strain, whereas all the cKP strains used in this study, were virulent as they cause >50% mortality within 24 hours. The varying mortality rate between the strains showed that there needs a detailed investigation at the genomic level to understand the virulence nature of *K. pneumoniae*. A detailed exploration of the virulence characteristics is needed to treat the patients infected by *K. pneumoniae*.^47^ In this study, it was evident that hypermucoviscous hypervirulent strains of *K. pneumoniae* were highly virulent which needs to be identified early to circumvent the extremity of infection. Notably, the cKP strains were also virulent, which must be tracked for the acquisition of hypervirulent characteristics through the transfer of virulent plasmids and infections must be treated with effective antibiotics. On the other hand, it is the first study from India, which utilized *G. mellonella* as an infection model to study the pathogenicity of cKP and hvKP strains. Our study has certain limitations as we did not study in detail the colistin resistance mechanism at the molecular level and in the virulence study, we screened only the limited virulence genes which is related to this study.

## Conclusion

Our study showed the dissemination of colistin-resistant HMV hvKP in the clinical isolates collected from the diagnostic centres situated in Tamil Nadu. It is important to take action to avoid the spread of such deadly superbugs in the hospital as well as in the community as they are highly resistant and virulent. This study also found that there are no *mcr* genes in the colistin-resistant *K. pneumoniae* isolates which shows that there could be another resistance mechanism that is responsible for the ineffectiveness of colistin that must be identified. We also reported the emergence of colistin-resistant hvKP ST23 and ST86 strains in Tamil Nadu. *In vivo* study implies that the hmv-hvKP strains were highly pathogenic and must be monitored properly to avoid their spread. It is the responsibility of clinicians to report and create awareness about identifying such pathogens and they must be characterized thoroughly to curb their dissemination and reduce the mortality and morbidity caused by such deadly superbugs.

## Data Availability

All data produced in the present study are available upon reasonable request to the authors.

https://www.ncbi.nlm.nih.gov/bioproject/?term=PRJNA838492

## Acknowledgement

The authors would like to thank the Vellore Institute of Technology for supporting this research in the form of the ‘VIT SEED’ grant. We are thankful to Dr Prakash KV, Professor, All India Network Project on Soil Arthropod Pests, Department of Entomology, University of Agricultural Sciences, Gandhi Krishi, Vignana Kendra, Bengaluru, Karnataka, for providing the wax worms.

## Author’s contribution statement

**Thamaraiselvan Shanthini:** Conceptualization, Methodology, Formal analysis, Resources, Investigation, Writing-Original Draft. **Prasanth Manohar:** Validation, Investigation, Resources, Writing-Review and Editing. **Xiaoting Hua:** Validation, Data curation, Software, Visualization. **Sebastian Leptihn:** Resources, Writing-Review and Editing. **Nachimuthu Ramesh:** Conceptualization, Resources, Supervision, Project administration.

## Ethical Statement

This study was conducted as per the ethical guidelines issued by the Institutional Ethical Committee for the studies on Human subjects (IECH) VIT/IECH/004/Jan28.2015.

## Data availability

The genome sequence of five *K. pneumoniae* strains was deposited under the BioProject: PRJNA838492 and NCBI accession numbers for the isolates reported in this paper are; KP1: JAMOKB000000000, KP2: JAMOKC000000000, KP8: JAMOKD000000000, KP9: JAMOKE000000000 and KP12: JAMOKA000000000.

