## Supplemental material for "Detection of Hypervirulent *Klebsiella pneumoniae* from Clinical Samples in Tamil Nadu"

**Supplementary materials:**

**Supplementary tables:**

**Table S1: The list of primers used for screening mobile colistin resistance gene (*mcr*)**

| **Gene** | **Primer sequence (5’-3’)** | **Product size (bp)** |
| --- | --- | --- |
| *mcr-1* | F- AGTCCGTTTGTTCTTGTGGC | 320 |
|  | R- AGATCCTTGGTCTCGGCTTG |  |
| *mcr-1* | CLR5: F- CGGTCAGTCCGTTTGTTC | 309 |
|  | CLR5: R- CTTGGTCGGTCTGTAGGG |  |
| *mcr-2* | F- CAAGTGTTGGTCGCAGTT | 700 |
|  | R- TCTAGCCCGACAAGCATACC |  |
| *mcr-3* | F- AAATAAAAATTGTTCCGCTTATG | 900 |
|  | R- AATGGAGATCCCCGTTTTT |  |
| *mcr-4* | F- TCACTTTCATCACTGCGTTG | 1100 |
|  | R- TTGGTCCATGACTACCAATG |  |
| *mcr-5* | F- ATGCGGTTGTCTGCATTTATC | 1644 |
|  | R- TCATTGTGGTTGTCCTTTTCTG |  |

**Table S2:** **The list of primers used for screening integrase genes and gene cassette region**

| **Gene** | **Primer sequence (5’-3’)** | **Product size (bp)** |
| --- | --- | --- |
| *IntI1* | F- GGTCAAGGATCTGGATTTCG | 436 |
|  | R- ACATGCGTGTAAATCATCGTC |  |
| *IntI2* | F- CACGGATATGCGACAAAAAGG | 788 |
|  | R- TGTAGCAAACGAGTGACGAAATG |  |
| *IntI3* | F- AGTGGGTGGCGAATGAGTG | 600 |
|  | R- TGTTCTTGTATCGGCAGGTG |  |
| *5’CS* | F- GGCATCCAAGCAGCAAG | Variable |
| *3’CS* | R- AAGCAGACTTGACCTGA |  |

**Table S3: The list of primers used for screening capsular serotypes in *K. pneumoniae***

| **Gene** | **Primer sequence (5’-3’)** | **Product size (bp)** |
| --- | --- | --- |
| K1 | F- GGTGCTCTTTACATCATTGC | 1283 |
|  | R- GCAATGGCCATTTGCGTTAG |  |
| K2 | F- GACCCGATATTCAATACTTGACAGAG | 641 |
|  | R- CCTGAAGTAAAATCGTAAATAGATGGC |  |
| K5 | F- TGGTAGTGATGCTCGCGA | 280 |
|  | R- CCTGAACCCACCCCAATC |  |

**Table S4:** **The list of primers used for screening virulence genes in *K. pneumoniae***

| **Gene** | **Primer sequence (5’-3’)** | **Product size (bp)** |
| --- | --- | --- |
| *rmp A* | F- ACTGGGCTACCTCTGCTTCA | 535 |
|  | R- CTTGCATGAGCCATCTTTCA |  |
| *Aerobactin* | F- GCATAGGCGGATACGAACAT | 556 |
|  | R- CACAGGGCAATTGCTTACCT |  |
| *KfuBC* | F- GAAGTGACGCTGTTTCTGGC | 797 |
|  | R- TTTCGTGTGGCCAGTGACTC |  |

**Table S5: The list of *K. pneumoniae* isolates and their clinical source.**

| **S. No.** | **Isolate ID** | **Bacteria** | **Location** | **Source** |
| --- | --- | --- | --- | --- |
| 1. | KP 1 | *K. pneumoniae* | Trichy | Sputum |
| 2. | KP 2 | *K. pneumoniae* | Madurai | Urine |
| 3. | KP 3 | *K. pneumoniae* | Trichy | Pus |
| 4. | KP 4 | *K. pneumoniae* | Chennai | Urine |
| 5. | KP 5 | *K. pneumoniae* | Chennai | Wound swab |
| 6. | KP 6 | *K. pneumoniae* | Chennai | Urine |
| 7. | KP 7 | *K. pneumoniae* | Chennai | Urine |
| 8. | KP 8 | *K. pneumoniae* | Trichy | Sputum |
| 9. | KP 9 | *K. pneumoniae* | Chennai | Blood |
| 10. | KP 10 | *K. pneumoniae* | Madurai | Urine |
| 11. | KP 11 | *K. pneumoniae* | Chennai | Blood |
| 12. | KP 12 | *K. pneumoniae* | Madurai | Urine |
| 13. | KP 13 | *K. pneumoniae* | Trichy | Blood |
| 14. | KP 14 | *K. pneumoniae* | Chennai | Urine |
| 15. | KP 15 | *K. pneumoniae* | Trichy | Pus |
| 16. | KP 16 | *K. pneumoniae* | Madurai | Urine |
| 17. | KP 17 | *K. pneumoniae* | Trichy | Pus |
| 18. | KP 18 | *K. pneumoniae* | Chennai | Pus |
| 19. | KP 19 | *K. pneumoniae* | Chennai | Urine |
| 20. | KP 20 | *K. pneumoniae* | Chennai | Urine |
| 21. | KP 21 | *K. pneumoniae* | Chennai | Urine |
| 22. | KP 22 | *K. pneumoniae* | Madurai | Sputum |
| 23. | KP 23 | *K. pneumoniae* | Chennai | Abdominal drain |
| 24. | KP 24 | *K. pneumoniae* | Chennai | Urine |
| 25. | KP 25 | *K. pneumoniae* | Chennai | Urine |
| 26. | KP 26 | *K. pneumoniae* | Madurai | Urine |
| 27. | KP 27 | *K. pneumoniae* | Trichy | Sputum |
| 28. | KP 28 | *K. pneumoniae* | Trichy | Urine |
| 29. | KP 29 | *K. pneumoniae* | Trichy | Urine |
| 30. | KP 30 | *K. pneumoniae* | Trichy | Sputum |

**Supplementary figure:**

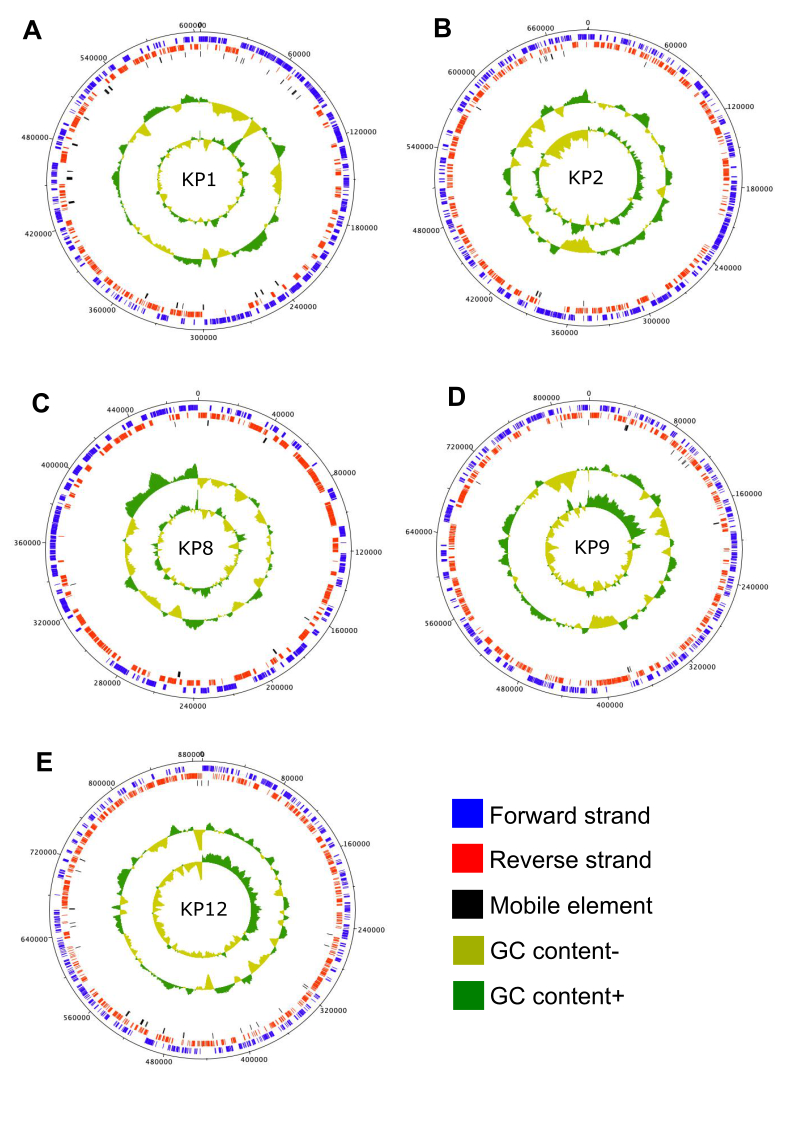

**Figure S1: The genome map of hypervirulent *Klebsiella pneumoniae* strains (A) KP1, (B) KP2, (3) KP8, (4) KP9, and (5) KP12.** Circles from inner to outer represent; circle 1. GC skew in olive (-) and green (+), 2. G+C content and deviation from the average, olive-below average and green-above average, 3. Mobile elements (black), 4. Position of CDS in the minus strand, 5. Position of CDS in the plus strand and 6. Scale (bp).
